# Fine-tuned Forecasting Techniques for COVID-19 Prediction in India

**DOI:** 10.1101/2020.08.10.20167247

**Authors:** Abhinav Gola, Ravi Kumar Arya, Animesh, Ravi Dugh, Zuber Khan

## Abstract

Estimation of statistical quantities plays a cardinal role in handling of convoluted situations such as COVID-19 pandemic and forecasting the number of affected people and fatalities is a major component for such estimations. Past researches have shown that simplistic numerical models fare much better than the complex stochastic and regression-based models when predicting for countries such as India, United States and Brazil where there is no indication of a peak anytime soon. In this research work, we present two models which give most accurate results when compared with other forecasting techniques. We performed both short-term and long-term forecasting based on these models and present the results for two discrete durations.

## 1. Introduction

In December 2019, some people in Wuhan, China were infected by the novel coronavirus, named 2019-nCoV and since then, this outbreak has spread to more than 200 countries all over the world. This has led the World Health Organization (WHO) to declare it as international public health emergency. Governments of the nations affected by this pandemic are running around to formulate provisions and provide resources to handle this epidemic. Forecasting the infection rate for a nation can act as a huge asset in planning and formulation of policies for such nations.

While no model can accurately forecast the rates of infection and mortality, attempts have been made to consider and analyse the strengths and shortcomings of many studies and models presented regarding the coronavirus. Whereas the forecast models used by the health department or Government of India were not disclosed, we can definitely continue with existing models in separate publications. Each of these models took different approaches and techniques to predict the future rates.

There has been a profusion of available mathematical techniques to predict the infection rate for the currently ongoing Covid-19 crisis. In past research [18], researchers evaluated the performance of majority of these techniques and concluded with two models which can be used for further purposes of estimating the number of cases affected by the coronavirus as these models gave the best predictions. These two models, exponential curve fitting and least square fitted model, can be used for short-term and long-term forecasting respectively.

In this study, we implement these two techniques on an updated dataset taken from the official website of Ministry of Health and Family Welfare, Government of India [17]. We estimate the number of affected, death and recovered cases for 2 different durations - one from August 5 to September 3 i.e. for 4 weeks, and the other from August 5 to September 23 i.e. for 7 weeks. We believe this forecast would assist the government and certain other official authorities in preparing and organizing necessary resources to deal with this pandemic.

This study is organized into five main sections. The paper starts with the general information about history and information of the disease. Section 2 provides the survey of the previously employed forecasting models to predict the confirmed cases in Indian context. We present our methodology in section 3 and discuss our findings and results in section 4. We conclude this research work in section 5 alongside providing scope for future improvements.

## 2. Related Work

Research on estimation of infection rate of Covid-19 has been quite prolific. Majority of these revolve around traditional machine learning methods and neural network-based models. R. Sujath et. al [11] and Ajit Kumar Pasayat et. al [16] used linear regression models while Gaurav Pandey et, al [12] employed polynomial regression technique to predict the Coronavirus cases in its early months. R. Sujath et. al [11] also used multi-layer perceptron models alongside their stochastic vector autoregression (VAR) time-series model. Another case of using complex learning models is Anuradha Tomar et. al [14] applying a LSTM model to forecast the number of cases.

In case of small epidemics, Meyers [1] studied the forecasted spread using a model of the Susceptible-Infected-Recuperated (SIR). In the simulation COVID-19 diffusion experiments, Wu et. al [2] applied the Susceptible-Exposed-Infectious-Recovered (SEIR) Model. Anastassopoulou Al. [3] performed a simulation study of situation COVID-19 at the very initial stage of pandemics, a model of susceptible-infectious-recovered-dead (SIRD) was used. Ghosh et al [4] used a pandemic model of Susceptible Infectious Susceptible (SIS) to forecast spread of the COVID-19 in India.

Kumar et. al [5], in order to analyse the Indian scenario, has used the ARIMA time series analysis technique. Their predictions were very similar to the later reported actual values. Basu [8] has been researching time-based viral spread in India on his own basis. According to his predictions, in early June, total number of cases in India was estimated to cross 200,000 and that prediction was quite accurate. Sudip Ghosh et. al [20] used linear square fitted modelling while Hemanta Kumar Baruah et. al [19] fitted an exponential curve for their predictions.

## 3. Methodology

### 3.1 Short-term forecasting [Exponential Curve fitting]

Short-term forecasting can be done based on elementary analytical approaches instead of diving into complex architectures like disease modelling or neural networks. Previous research [18] has shown that for shorter durations, simplistic curve fitting models achieve better accuracy than regression and pandemic models. Observing the patterns of number cases in countries such as China, Spain and Italy we can infer that the natural infection rate curve will follow a non-linear path initially till it hits its peak and begins to subside. Nations such as India, United States of America and Brazil are still in the nonlinear portion of the plot and due to the uncertainty of their peak point, forecasting for such countries can only be done for short durations.

**Figure 1.**
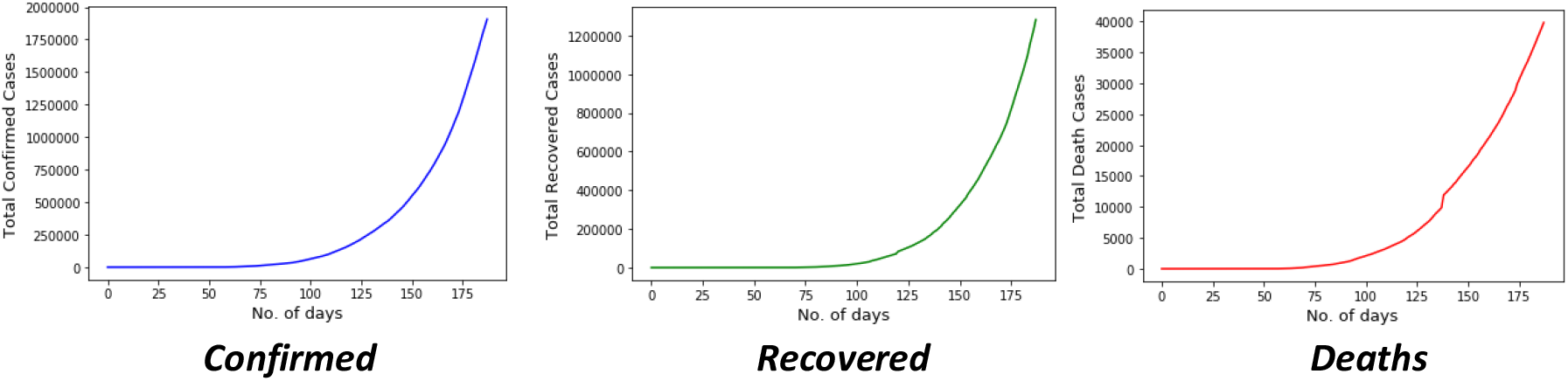
Plots of cumulative cases in India.

Observing the pattern for India, we can discern a definite exponential trajectory, which can be exploited by studying the time series data in an inverted fashion and then instituting a numerical model established on the latter part of the data. Let *P(t)* be the total number of affected cases. *Q(t)* be the total number of death cases, and *R(t)* be the total number of recovered cases at a given time t. To verify out assumption of the curve being exponential, we took the natural logarithm of *P(t), Q(t)* and *R(t)*. The resulting plots shown in Fig. 2 are linear for each curve, thus establishing our assumption as legitimate.

**Figure 2.**
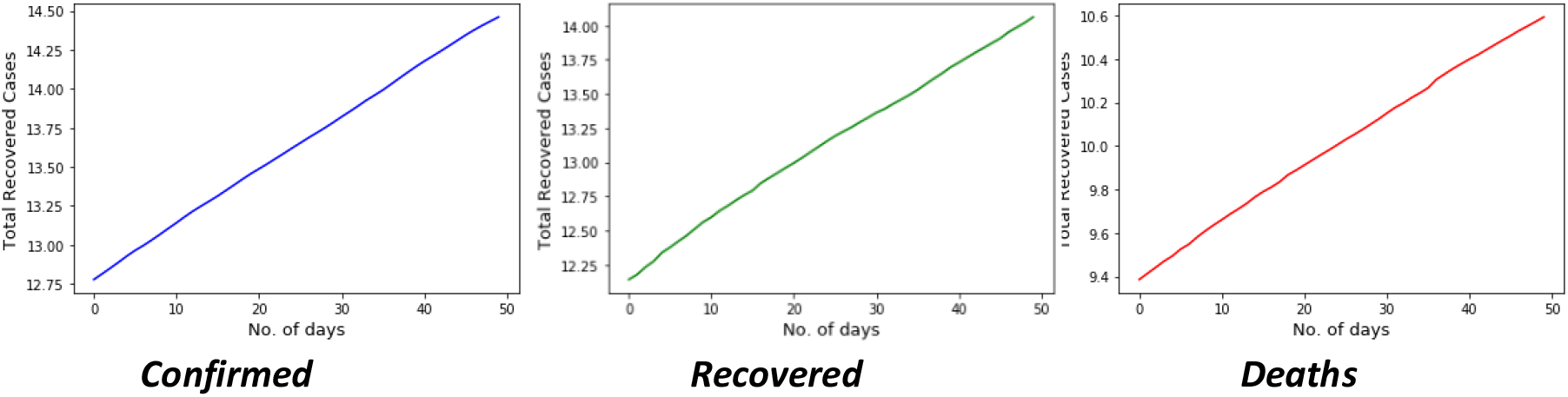
Plots of natural logarithm of cumulative cases in India.

Fitting against exponential functions is exceedingly fragile because tiny variations in the exponent can result in large differences in the result. Optimising is done across many orders of magnitude, and errors near the origin are not equally weighted compared to errors higher up the curve. The simplest way to handle this is to convert our exponential data to a linear form using a natural logarithm transformation: Considering the equations of curve to be:

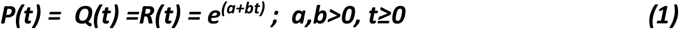

where a and b are constants. Taking natural logarithm of both sides:

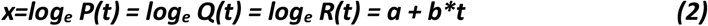

This allows us to use the linear curve fitting method instead of the slower polynomial fitting method which when employed on large values is prone to result in overflow errors. We would later transform the data back into linear space for analysis. We used the *polyfit()* function of the *numpy* module placed in *Python* and got the coefficients’ values as:

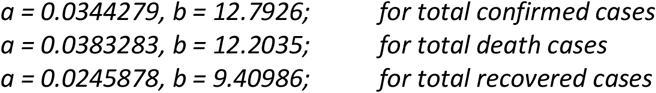

rendering our equations to be:

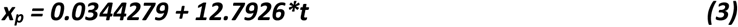

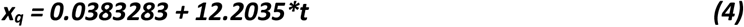

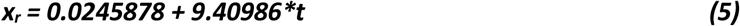

The covariance matrices obtained for each case are shown in Fig. 3.

**Figure 3.**
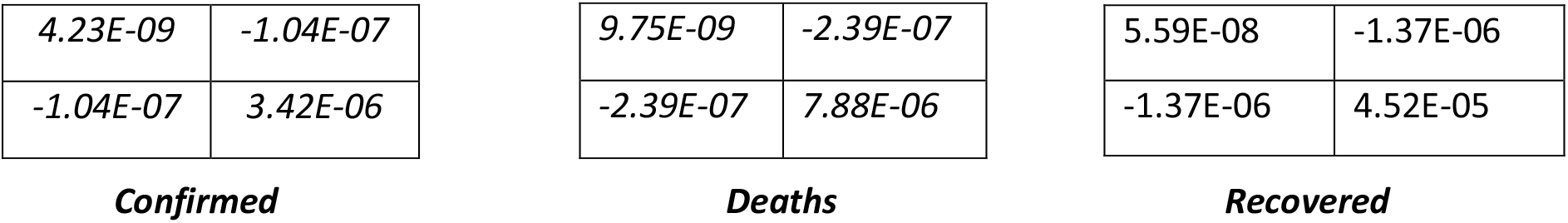
Covariance matrices after curve fitting.

### 3.2 Least square fitting method

A methodology widely used to perform regression analysis is the least square regression method. This is a statistical technique to determine the best line of fit between an independent and a dependent variable. The ‘least-square method’ combines measurements in order to extract the parameter estimates that define the curve that best matches the results. Using the least square rule, given the set of N (noisy) measurements f_i_, i∈1, N, which are to be applied to the curve f(a), where ‘a’ is the vector of the parameter values, we seek to minimize the square of the difference between the measurements and the values of the curve to provide an approximation of the parameters ‘a^∧^’ according to ***(7)***

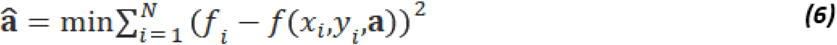

When we fit our data to the polynomial function graph, the polynomial curve fit is. The same technique of smallest squares is used to identify a certain degree polynomial which has a minimum overall error:

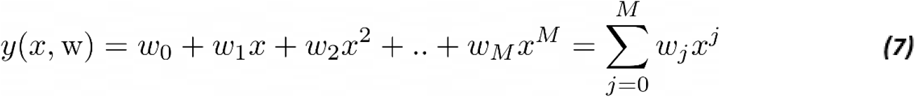

where M is the order of the polynomial

We obtain a fit by minimizing an error function – sum of squares of the errors between the

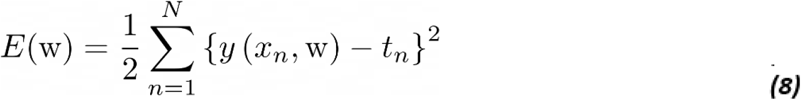

predictions y(x_n_,w) for each data point x_n_ and target value t_n_.

Here, polynomial of degree 6 is used for fitting the dataset.

## 4. Results

Observing the non-linear pattern in India’s COVID-19 infection rate, we employed curve fitting techniques to predict the number of confirmed, death and recovered cases for both short-term and long-term durations. Due to the unpredictable nature of the exponential graph, small modifications in input can lead to abrupt changes in our output. Thus, we used exponential curve fitting for short-term forecasting for a duration of 4 weeks starting from August 8, 2020 to September 4, 2020. Polynomial regression modelling is used for long-term forecasting for a duration of 7 weeks starting from August 8, 2020 to September 24, 2020.

**Table 1.** Forecasted cases from August 5 to August 17 [Exponential Curve Fitting].

| Dates | Forecasted Confirmed Cases | Forecasted Death cases | Forecasted Recovered Cases |
| --- | --- | --- | --- |
| 08-08-2020 | 22,30,000 | 44,937 | 15,20,000 |
| 09-08-2020 | 23,10,000 | 46,055 | 15,80,000 |
| 10-08-2020 | 23,90,000 | 47,202 | 16,40,000 |
| 11-08-2020 | 24,70,000 | 48,377 | 17,10,000 |
| 12-08-2020 | 25,60,000 | 49,581 | 17,70,000 |
| 13-08-2020 | 26,50,000 | 50,815 | 18,40,000 |
| 14-08-2020 | 27,40,000 | 52,080 | 19,10,000 |
| 15-08-2020 | 28,40,000 | 53,377 | 19,90,000 |
| 16-08-2020 | 29,40,000 | 54,705 | 20,70,000 |
| 17-08-2020 | 30,40,000 | 56,067 | 21,50,000 |
| 18-08-2020 | 31,50,000 | 57,463 | 22,30,000 |
| 19-08-2020 | 32,60,000 | 58,893 | 23,20,000 |
| 20-08-2020 | 33,70,000 | 60,359 | 24,10,000 |

Results for each case are presented in Tables 1 and 2 while their respective plots are demonstrated in Figs. 5 and 6. As per our forecasts, the total number of cases in India would cross 30,00,000 by August 15, 2020. By August 25, 2020 it would cross 40,00,000, and around September 1, it should exceed the 50,00,000 value.

**Figure 4.**
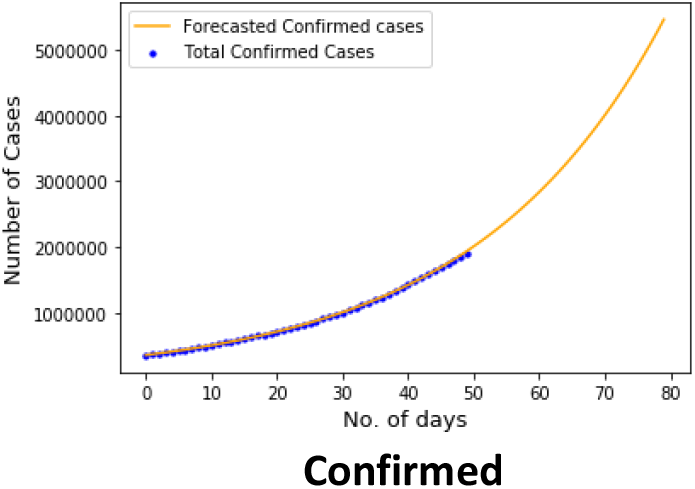
Forecasting plots for exponential curve fitting.

**Table 2.** Forecasted cases from August 17 to September 3 [Exponential Curve Fitting].

| **Dates** | **Forecasted Confirmed Cases** | **Forecasted Death cases** | **Forecasted Recovered Cases** |
| --- | --- | --- | --- |
| 21-08-2020 | 34,90,000 | 61,861 | 25,00,000 |
| 22-08-2020 | 36,10,000 | 63,401 | 26,00,000 |
| 23-08-2020 | 37,40,000 | 64,980 | 27,00,000 |
| 24-08-2020 | 38,70,000 | 66,597 | 28,10,000 |
| 25-08-2020 | 40,00,000 | 68,255 | 29,20,000 |
| 26-08-2020 | 41,40,000 | 69,954 | 30,30,000 |
| 27-08-2020 | 42,90,000 | 71,695 | 31,50,000 |
| 28-08-2020 | 44,40,000 | 73,480 | 32,70,000 |
| 29-08-2020 | 45,90,000 | 75,309 | 34,00,000 |
| 30-08-2020 | 47,50,000 | 77,184 | 35,30,000 |
| 31-08-2020 | 49,20,000 | 79,105 | 36,70,000 |
| 01-09-2020 | 50,90,000 | 81,074 | 38,20,000 |
| 02-09-2020 | 52,70,000 | 83,092 | 39,70,000 |
| 03-09-2020 | 54,60,000 | 85,161 | 41,20,000 |
| 04-09-2020 | 56,47,890 | 87,280 | 42,81,130 |

**Figure 5.**
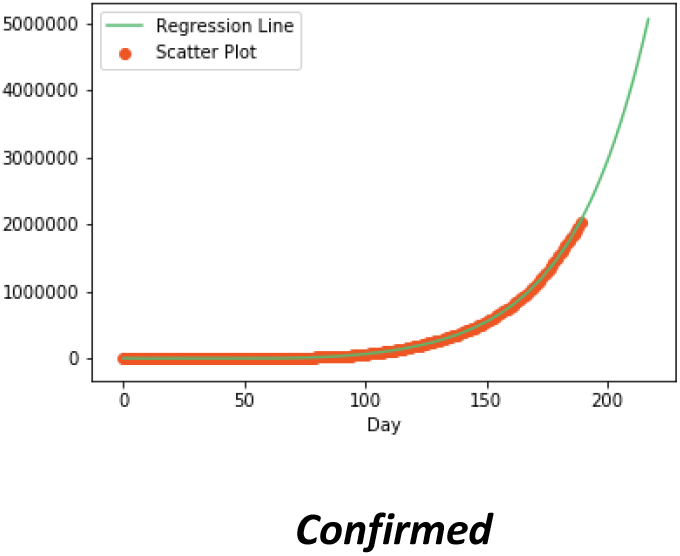
Forecasting plots for least squared error fitting.

**Table 3.** Forecasting from August 8, 2020 to August 17, 2020 [Least Squared Error Fitting].

| **Date** | **Total Confirmed** | **Total Deceased** | **Total Recovered** |
| --- | --- | --- | --- |
| 08-08-20 | 2189023 | 43040 | 1467800 |
| 09-08-20 | 2262220 | 43897 | 1520544 |
| 10-08-20 | 2337755 | 44765 | 1575053 |
| 11-08-20 | 2415696 | 45643 | 1631383 |
| 12-08-20 | 2496114 | 46531 | 1689587 |
| 13-08-20 | 2579082 | 47430 | 1749723 |
| 14-08-20 | 2664673 | 48339 | 1811849 |
| 15-08-20 | 2752963 | 49259 | 1876024 |
| 16-08-20 | 2844029 | 50190 | 1942310 |
| 17-08-20 | 2937950 | 51132 | 2010770 |

**Table 4.**
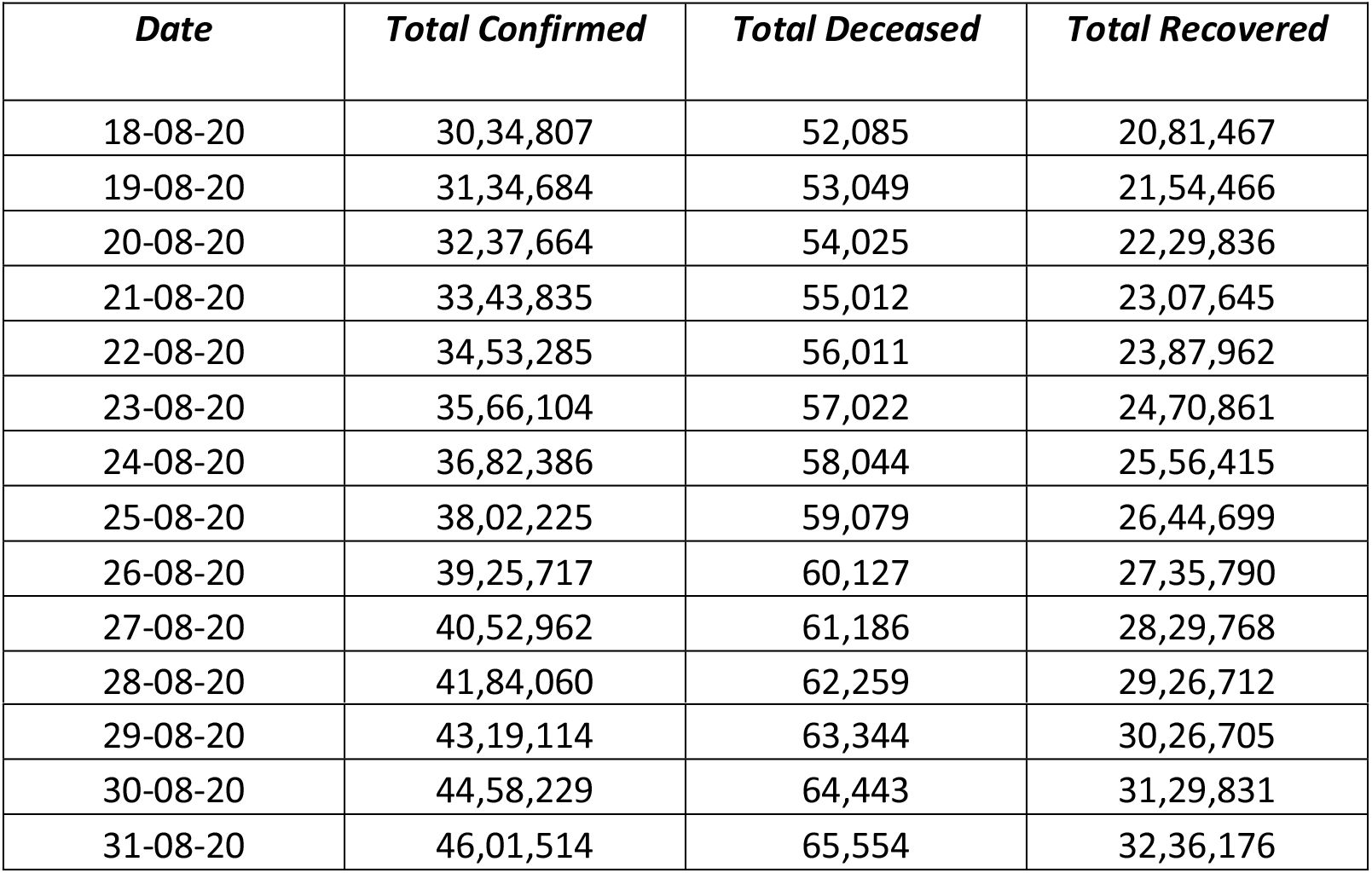
Forecasting from August 18, 2020 to August 31, 2020 [Least Squared Error Fitting].

**Table 5.** Forecasting from 1 September 2020 to 24 September 2020 [Least Squared Error Fitting].

| *Date* | *Total Confirmed* | *Total Deceased* | *Total Recovered* |
| --- | --- | --- | --- |
| 01-09-20 | 4749077 | 66680 | 3345829 |
| 02-09-20 | 4901031 | 67820 | 3458879 |
| 03-09-20 | 5057489 | 68973 | 3575417 |
| 04-09-20 | 5218569 | 70141 | 3695539 |
| 05-09-20 | 5384390 | 71324 | 3819338 |
| 06-09-20 | 5555073 | 72521 | 3946913 |
| 07-09-20 | 5730742 | 73733 | 4078365 |
| 08-09-20 | 5911523 | 74961 | 4213794 |
| 09-09-20 | 6097545 | 76205 | 4353304 |
| 10-09-20 | 6288940 | 77465 | 4497002 |
| 11-09-20 | 6485841 | 78741 | 4644997 |
| 12-09-20 | 6688386 | 80034 | 4797398 |
| 13-09-20 | 6896713 | 81345 | 4954318 |
| 14-09-20 | 7110964 | 82672 | 5115873 |
| 15-09-20 | 7331285 | 84018 | 5282179 |
| 16-09-20 | 7557823 | 85381 | 5453356 |
| 17-09-20 | 7790727 | 86764 | 5629526 |
| 18-09-20 | 8030151 | 88165 | 5810813 |
| 19-09-20 | 8276251 | 89586 | 5997345 |
| 20-09-20 | 8529185 | 91027 | 6189250 |
| 21-09-20 | 8789116 | 92488 | 6386659 |
| 22-09-20 | 9056207 | 93971 | 6589708 |
| 23-09-20 | 9330628 | 95474 | 6798532 |
| 24-09-20 | 9612548 | 96999 | 7013271 |

## 5. Conclusion

Building upon the previous research [18], current study implemented two numerical models to forecast the number of cases related to COVID-19 in India, namely – exponential curve fitting and least square fitted model. Both of the models forecasted an upward of 30 lakhs cases and 40,000 deaths for the upcoming months. Unless there is a sudden peak in the graph and it begins to subside, we are going to face an enormous challenge to handle this pandemic. To prevent a dearth of required resources, government and official organisations should plan factoring in the forecasted cases.

This study can be expanded to establish other mathematical and regression techniques for the forecasting of the COVID-19 cases in future. This would be essential in having a diverse assortment of prediction techniques to consider while developing new policies.

## Data Availability

N/A

